# The effects of Far-UVC irradiation on the presence and concentration of ESKAPEE pathogens on hospital surfaces: study protocol for a multi-site, double-blinded randomized controlled trial in La Paz, Bolivia

**DOI:** 10.64898/2026.02.04.26345557

**Authors:** Lindsay B. Saber, Melani Rojas, Darcy M. Anderson, Deverick J. Anderson, Holger Claus, Ryan Cronk, Karl G. Linden, Megan E.J. Lott, Lewis J. Radonovich, Bobby G. Warren, Richard D. Williamson, Richard L. Vincent, Sergio Gutiérrez-Cortez, Carla Calderón Toledo, Joe Brown

## Abstract

Hospital-acquired infections are a known and growing problem worldwide. Far-UVC is a novel disinfection method that inactivates bacteria with limited penetration into human skin or eyes. A clustered, unmatched, randomized control trial (RCT) will be implemented in two Bolivian hospitals. The intervention arm will receive functioning Far-UVC lamps, whereas the control arm will receive identical lamps that do not emit UV light (shams). Based on baseline data, 40 lamp fixtures will be installed above hospital sinks, 10 per arm per hospital. Environmental samples (air and surface swabs) will be collected and analyzed via culture and sequencing. Simultaneously, air chemical monitoring data will be collected.

## Introduction

Healthcare-acquired infections (HAIs) are a global problem. The CDC estimates that one of every 31 hospitalized patients in the United States contracts an HAI (1). The burden is demonstrated to be much higher in low- and middle-income countries (LMICs), where lack of consistent antimicrobial stewardship has contributed to the rapid emergence of resistant bacteria(2). The leading cause of HAIs worldwide are ESKAPEE pathogens (*Enterococcus faecium*, *Staphylococcus aureus*, *Klebsiella pneumoniae*, *Acinetobacter baumannii*, *Pseudomonas aeruginosa*, *Enterobacter* spp., and *Escherichia coli*) (3). ESKAPEE pathogens have the potential to develop multidrug resistance.

Premise plumbing is increasingly recognized as an important factor in HAIs. Sink faucets, basins, and drains can develop biofilms containing diverse microbes, including pathogens and AMR genes (4, 5). The transmission of pathogens between sink drains and patients is bi-directional, with evidence suggesting that patient-derived pathogens can quickly appear in sink drains (6), that sink drains can be colonized via liquid disposal down sinks (7), and that drains can subsequently present microbial risks to patients(8). Several outbreaks have been attributed to sinks (9), primarily reported in the last three years (5, 6, 10–21); many implicated bacteria are antibiotic- and disinfectant-resistant.

Mitigation strategies have had variable success(22–24). These include chemical disinfectant treatments(23, 25, 26), mechanical cleaning(6), vibration and heat(22) or heat combined with disinfectant(27), bacteriophage treatment(28), and other measures, through recolonization of sinks and even horizontal transmission between sinks and drains in a plumbing network(29, 30) has been shown to occur rapidly, and repeated treatments are usually necessary once a drain network has been colonized.

Other well-characterized methods for reducing biofilm-associated risks – like the addition of bacteriostatic or biocidal metals (copper, silver, selenium compounds)(31), quorum sensing(32), and probiotic treatment (e.g., with *Bacillus*)(33) have been proposed but have not been widely evaluated. Sink replacement, drain replacement, and point-of-use filtration have been proposed but are not considered effective long-term strategies; maintaining sterility is also not feasible(34). New facilities can be colonized quickly(6) and a quick response to known colonization may be key(6) in excluding pathogens from drain networks, but repeated cleaning is usually needed, and even that may not be effective over time(24).

Studies have examined the sink “splash zone” and found increased detection of microbial targets near sinks and drains(35). It is suspected that a two-meter distance around sinks is susceptible to direct sink contamination via droplets ejected from p-traps, the bend in a sink drainpipe which holds a small amount of water, though the origin of this radius is unclear. Aerosol ejection from sinks is poorly characterized. Most studies of these phenomena have been conducted in high-income countries. However, hospital environments in low- and middle-income countries may experience higher microbial loads, representing a high potential for infection risk (36).

Far-UVC, an innovative disinfection technology, is expected to reduce pathogen concentrations significantly and therefore may help prevent outbreaks of ESKAPE pathogens (37, 38). The efficacy of Far-UVC in inactivating pathogens is well characterized in laboratory settings, though few studies have reported on *in situ* results (39–47). The efficacy of Far-UVC in eliminating ESKAPEE pathogens in hospital environments remains poorly studied.

## Materials and Methods

In this study, Far-UVC lamps will be mounted above certain sink areas in a hospital setting. The rooms in which the lamps will be installed serve a variety of uses, but none are explicitly inpatient rooms. Patient beds are not present, and doctors will not see patients in these rooms. However, patients, visitors, and other non-healthcare individuals may be in the room with a lamp for a limited time (example: a bathroom). To be eligible for inclusion, sinks must be positioned such that non-healthcare staff would not be exposed to the lamp installed above the sink for more than one hour per day. Therefore, those who occupy the room will either be standing or sitting, but never laying down. Accurate predictions of light distribution within a space are easily modeled, and therefore, the exposure to 222 nm light is predictable. Below is an example of a lamp installed above a sink. Four scenarios are depicted: an individual standing still directly below the lamp for 2 hours; an individual either sitting or standing 1 m away from the sink for 8 hours; and an individual sitting 2 m away from the sink for 8 hours. All these scenarios are extremely conservative and unlikely, as staff do not spend much time near sinks during their shifts. However, even in these unlikely scenarios, the modeled exposure limits remain well below ACGIH guidelines, never exceeding 8.4% of the recommended tolerable dose. The table below indicates the total hours required to remain in each position to reach the 8-hour workday limit.

**Figure 1.**
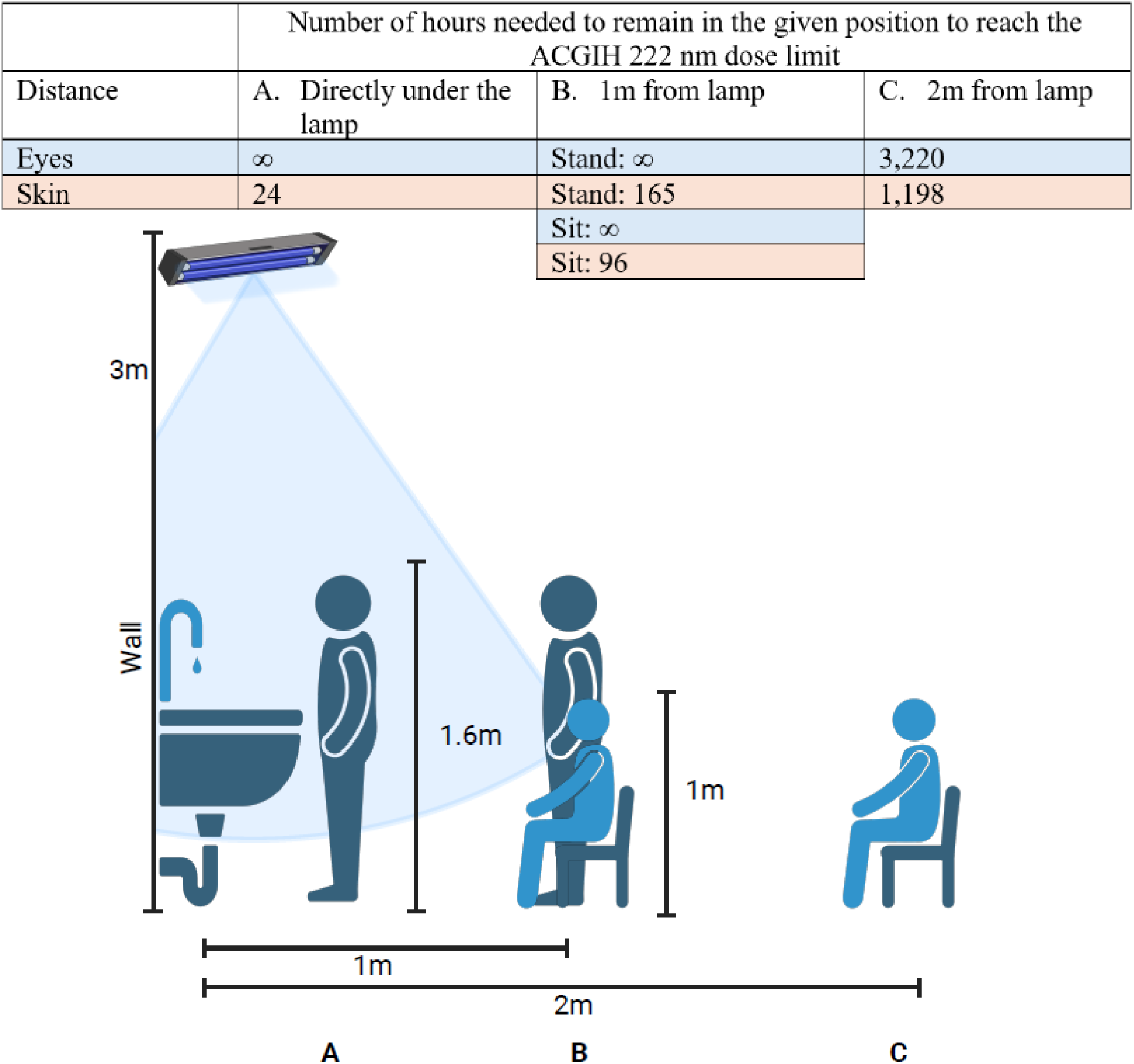
Number of hours needed to be spent either sitting or standing at distances A (0m), B (1m), or C (2m) away from the mounted Far-UVC lamp to be exposed to the ACGIH 222nm dose limit. It is assumed that the lamp is mounted 3m in height, and the individual is 1.6m standing and 1m sitting.

The above modeling clearly shows that the expected exposure, even under a very long, unlikely assumed residence time of people in the vicinity of lamps, will be minimal relative to the lower than allowable ACGIH exposure limits. Through evidence presented comprises substantial data to support the safety of the Far-UVC hospital trial.

### Setting

In Bolivia, antibiotics are commonly prescribed but also available without prescriptions; therefore, the barrier to receiving them is low, and dosages are highly variable. HAIs are common, especially when open wounds are exposed. Continuous disinfection of the air and surfaces is difficult, as HVAC systems are uncommon in this region of Bolivia, and bleach and other chemical disinfectants are largely ineffective in practice for eliminating ESKAPE pathogens from premises plumbing(48). The air and surfaces around sinks are especially contaminated with pathogens from the p-trap(49).

The La Paz is Bolivia’s second largest city and one of the world’s highest major cities, at an elevation of approximately 12,000 feet. Its neighboring city, El Alto, is even higher at 13,000 feet. One hospital, Hospital B, is a public, higher-resourced hospital in La Paz’s city center run by the National Health Fund. The other, Hospital A, is a municipal hospital in El Alto that primarily serves patients of limited means.

The two hospitals in La Paz could benefit substantially from Far-UVC. It is feasible that a measurable difference in ESKAPEE pathogens in the environment could be detectable, given their high baseline concentrations. Far UV has been proven to work on handwashing basins in a laboratory setting (37, 50). Given the high pathogenic risk posed by hospital sinks in Bolivia, a clear next step is to examine whether Far-UVC reduces pathogen levels in these environments.

### Previous Work

Laboratory studies to assess the efficacy of Far-UVC against ESKAPEE pathogens on typical hospital surfaces are underway. The goal of these studies is to determine the inactivation rates for each ESKAPEE pathogen on the various surfaces; data that does not exist in the literature.

Baseline sampling was conducted from May 2024 to May 2025 across various sinks within Hospital A and Hospital B. Environmental samples (air, surface, p-trap, and tap water) were collected in May-July 2024. These samples were transported back to UNC for multiplex qPCR analysis via TaqMan Array Cards and 16S amplicon sequencing. Additional air and surface samples were collected in February and May 2025 for culture and 16S amplicon sequencing. Whole genome sequencing was also performed on culture isolates.

Sample size calculations for the intervention are based on culture results from the 198 air and surface samples collected in May 2025. Each sample was plated on selective CHROMagar media (ESBL,

*Acinetobacter*, *Pseudomonas*, Orientation) and Tryptic Soy Agar to determine the presence or absence of ESKAPEE pathogens. The results confirmed a high prevalence of ESKAPEE pathogens, with 75% of Hospital B and 85.1% of Hospital A testing positive. A further breakdown of these data is presented in Table 1.

**Table 1.**
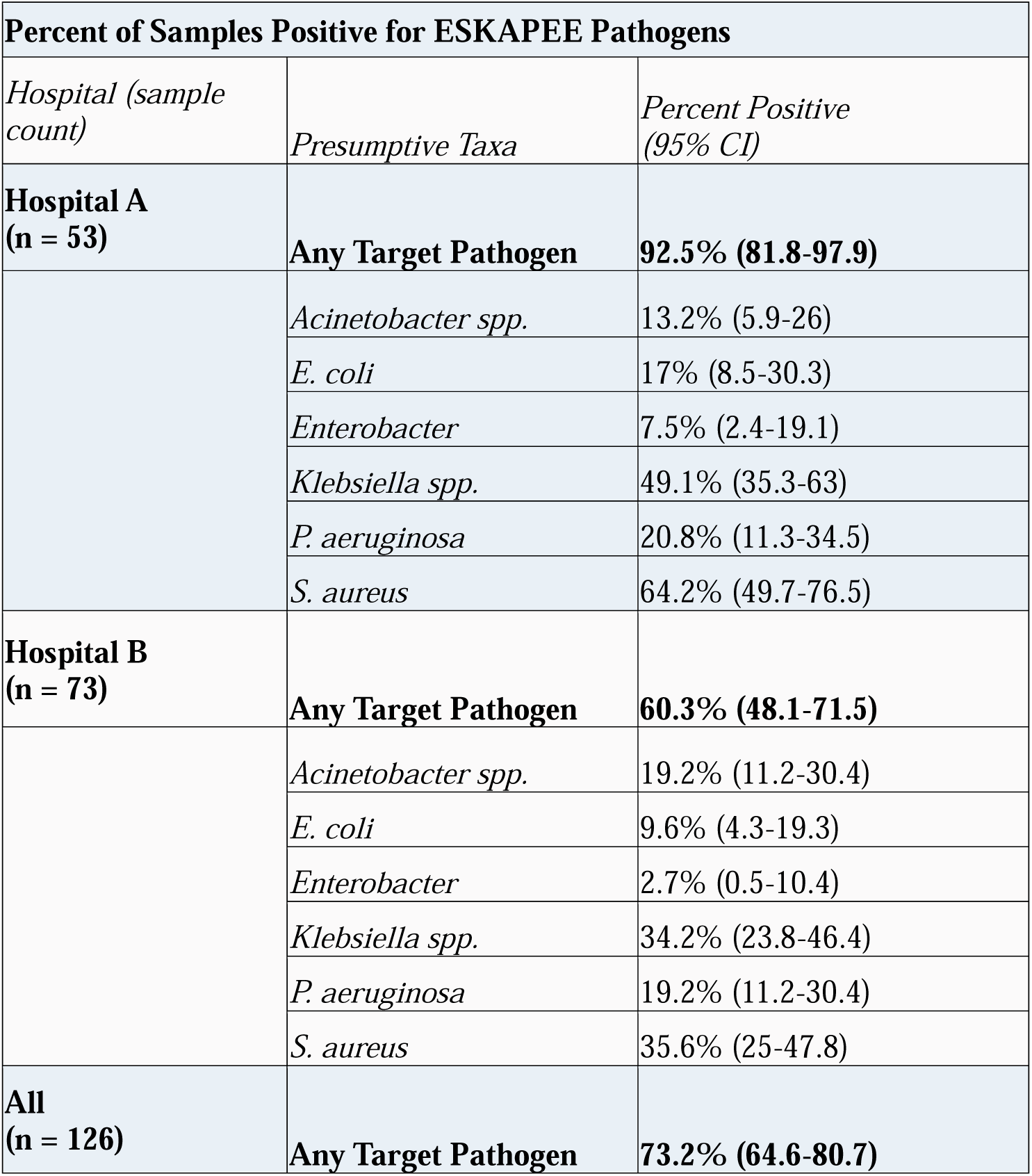
From these results, a baseline prevalence of ESKAPEE pathogens in the targeted areas has been characterized, and sample size calculations are discussed below.

### Trial overview

Our goal is to evaluate the efficacy of Far-UVC on hospital sink surfaces. The intervention will include installing 222nm lamps above sinks and collecting environmental samples to assess the prevalence of ESKAPEE pathogens. This protocol details a double-blind, RCT of the implementation of a targeted Far-UVC lamp in two Bolivian hospitals.

Inclusion and exclusion criteria are evaluated on the sink level, based on accessibility, functionality, and room use, as outlined in the methods. For those included in the study, full measurements of the sink area will be taken, and A3 Lighting Consultants will conduct simulations of lamp placement and countertop dosage. Beams will target the sinks and the surrounding countertop area directly as potential sites of exposure. A total of 40 lamps will be installed, 20 per hospital (see sample size section for more details). Half of the lamps (20 total, 10 per hospital) will be functioning Far-UVC lamps, and the other half (20 total, 10 per hospital) will be sham lamps. The sample size is sufficient for standalone analyses at both facilities, as this is a multi-site parallel trial. Figure 2 illustrates a mock-up of the model and lamp placement.

**Figure 2.**
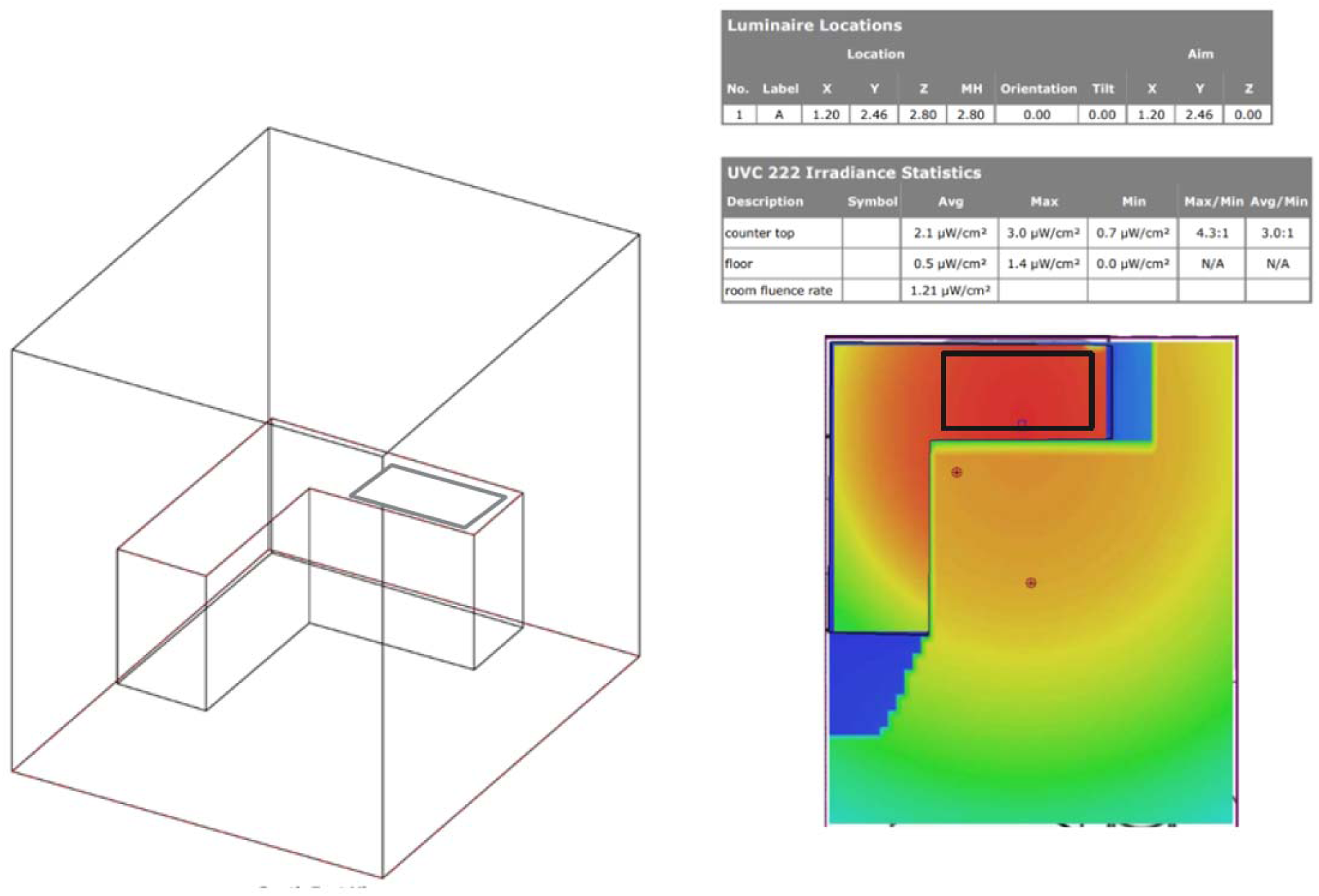
Example of sink model and lamp placement. Each sink included in the study will have a similar model built.

In half of the selected locations (20 sinks in total, 10 per hospital) it is planned to install placebo lamps. The sham (placebo) lamp will serve as a control. The sham lamps are visually indistinguishable from the intervention lamps but will not emit 222 nm irradiation. This is achieved by installing a filter on the lamps that blocks the germicidal light. The installation of sham lamps is intended to ensure that cleaning practices are consistent across intervention arms, as reactivity (in the form of changes in behavior or cleaning regimens) to a perceived intervention can introduce bias. Ushio Inc. B1 modules will be used in UVX mounting fixtures. Figure 2 outlines the overall study design, including the allocation of intervention and control sites.

**Figure 3.**
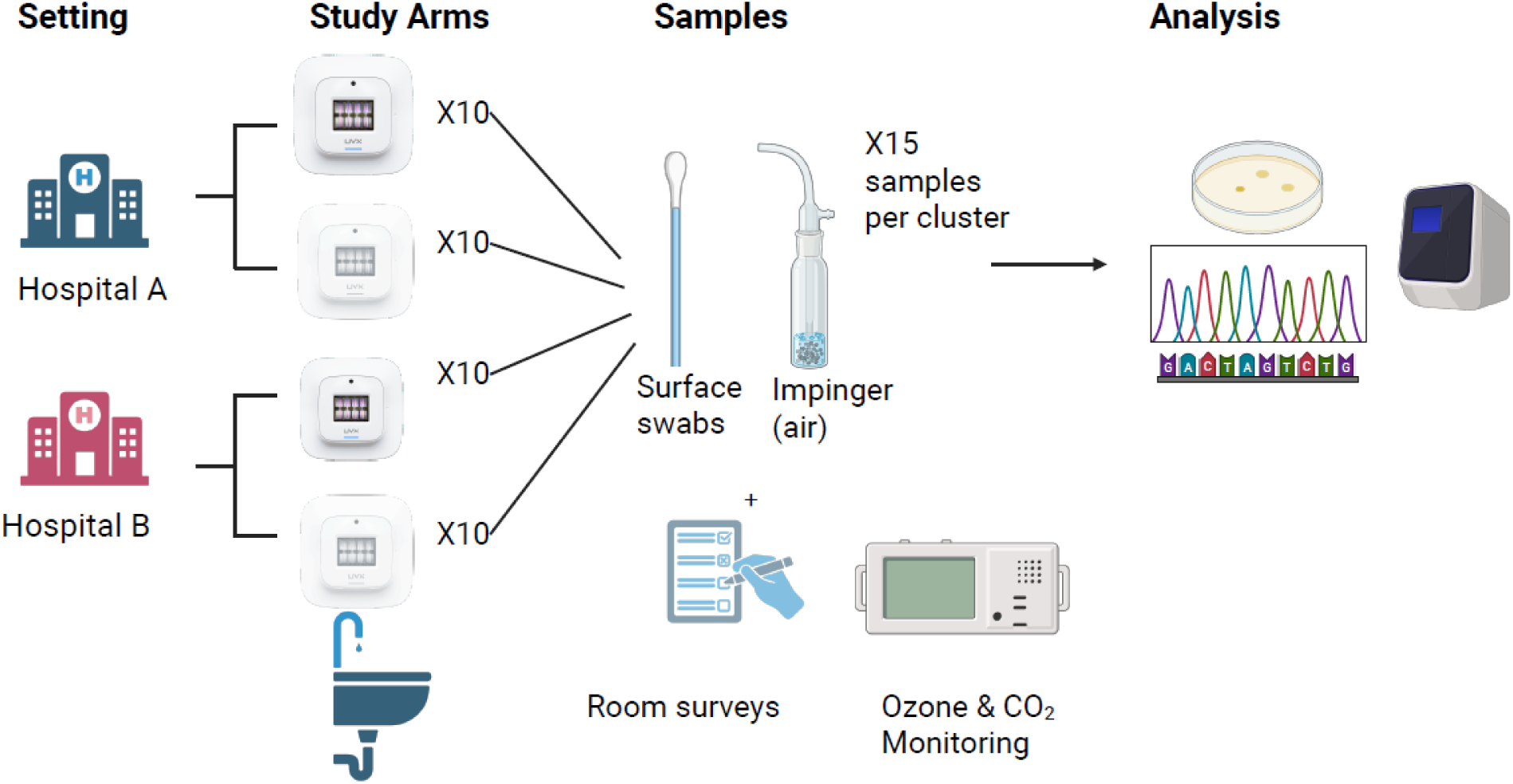
Overall study design, including samples to be collected.

Far-UVC is highly effective at inactivating bacteria in laboratory and some *in situ* settings, as noted in the introduction. We hypothesize the effectiveness will be transferred to clinical setting. The models created before the lamp installation will help us assess the theoretical UV dose delivered to each sink countertop. After lamp installation and at each sampling event, radiometers will be used to measure the irradiance received on surfaces. This will ensure consistency between locations and the functionality of each lamp. To ensure exposure levels are safe, the rooms will be designed so that a human of average height (1.6m) will never be exposed to more than the ACGIH guidelines of 479 mJ/cm^2^ for skin and 161 mJ/cm^2^ for eyes. Based on preliminary models, we estimate the sink surface dose will be approximately 190 mJ/cm2 on a 24-hour average at the counter. Based on these exposure levels it is expected to measure a statistically significant difference in ESKAPEE pathogen prevalence in areas where the 222 nm lamps are installed.

We will be monitoring for potential disinfection byproducts in the air, including ozone, PM2.5, PM10, formaldehyde, VOCs, and AQI, but laboratory studies suggest that these should not be of concern (51). Lamps will not be removed after the study concludes, at the hospital’s request; therefore, any benefits from Far-UVC use will continue. Filters will also be removed from shams so that both hospitals can use the full germicidal properties of all 20 lamps.

Below is the proposed protocol and data analysis plan for the Far-UVC intervention trial conducted in two Bolivian hospital test beds, funded by the Precision Microbiome Engineering Center (PreMiEr). This study was determined to be non-human-subject research by the UNC Internal Review Board (25–0455).

### Study outcomes, hypotheses, and approaches

This section outlines each of our study objectives and the accompanying hypothesis. Table 1 summarizes the primary, secondary, and tertiary objectives, with each explained in greater detail below. Methods are presented in this section but are expanded in the methods section.

### Primary Outcome: Does the intervention effect the prevalence of culturable ESKAPEE pathogens on surfaces (binary outcome)?

*Hypothesis*: The intervention reduces the prevalence of ESKAPEE pathogens on surfaces in proximity to the exposed sinks.

*Approach*: Once the lamps are installed and the initial waiting period is complete, a standard set of surface swabs will be collected from all sinks in both arms of the study. In the laboratory, samples will be plated onto selective media for ESKAPEE pathogens and incubated overnight. The results will be reported as a binary outcome: the presence or the absence of presumptive ESKAPEE pathogens. A regression analysis will be conducted with the blinded binary longitudinal data to compare the prevalence of any ESKAPEE pathogen detected, adjusting for clustering over time and by location.

### Secondary Outcome 1: Does the intervention effect the concentration of culturable ESKAPEE pathogens on surfaces (continuous outcome)?

*Hypothesis*: The intervention reduces the concentration of any ESKAPEE pathogen detected in proximity of the exposed sinks, with adjusting for clustering over time and by location.

*Approach*: The same samples from above will be used for secondary outcome 1. However, after overnight incubation, colonies of each presumptive ESKAPEE pathogen will be counted and reported as consistent with the normal distribution of continuous outcomes in culture CFUs per cm^2^. An analyst-blinded regression analysis will be used to compare the concentration of ESKAPEE pathogens between all sites, and if the intervention is associated with the difference.

### Secondary Outcome 2: Does the intervention effect the prevalence of ESKAPEE pathogens in aerosols?

*Hypothesis*: The intervention reduces the prevalence of ESKAPEE pathogens in aerosols in proximity of the exposed sinks.

*Approach*: For every set of surface swab samples collected, an air sample will be taken 1m away from the center of the sink, while cold water is running. Information about water pooling and drainage will be recorded. These samples will be plated on media selective for ESKAPEE pathogens, as in primary objective 1 and secondary objective 1. CFUs will be counted after overnight incubation. Regression analysis will be conducted with blinded data, to compare study arms, clustering over time and location.

### Secondary Outcome 3: Does the intervention have an effect on the microbial community composition?

*Hypothesis*: The intervention changes key measures of microbial community composition in proximity of the exposed sinks.

*Approach*: After the swab samples are plated for culture, the remaining buffer will be preserved and sent for metagenomic sequencing. Likely, the samples selected for sequencing will either be a representative subset of all samples, or we will pool based on similar sampling locations and/ or ward function. Regression analysis will be conducted to compare microbial communities in the metagenomic data between study arms.

### Secondary Outcome 4: Does the intervention affect predicted infections?

*Hypothesis*: The intervention reduces predicted ESKAPEE-associated infection risk in proximity of the exposed sinks.

*Approach*: A quantitative microbial risk assessment (QMRA) will be conducted using the quantitative, continuous culture data to model the risk of infection from ESKAPEE pathogens on hospital surfaces and in the air. Methods will mirror Capone et al. 2023 to predict ratio measures of effect between control and intervention arms (52).

### Secondary Outcome 5: Does the intervention effect the total microbial activity?

*Hypothesis*: The intervention reduces total microbial activity on hospital countertops in proximity of the exposed sinks.

*Approach*: A newer detection method for environmental microbial activity is fluorescence ATP monitors. Surface swabs are taken and immediately placed into a machine, which gives a fluorescence reading(53). At the time of sample collection, an additional ATP-specific surface swab will be taken directly adjacent to a countertop sample, within the bounds of a template(54). The swab will analyzed via Hygiena UltraSnap™ (Hygiena Camarillo, CA, USA), to measure the fluorescence of environmental ATP for a near-real-time measure of total metabolic activity in the microbial community. A regression analysis will compare environmental ATP concentrations between intervention and control arms.

### Secondary Outcome 6: Does the intervention effect ozone concentrations in the air?

*Hypothesis*: The intervention will increase indoor ozone concentrations but will not exceed 80 ppb, the ACGIH threshold in proximity of the exposed sinks.

*Approach*: Four ozone monitors, two in intervention and two in control rooms (for ozone background monitoring), will be installed 1.6 meters off the ground, average human height, on the wall adjacent to the mounted lamp (55). Continuous ozone monitoring data will be collected over the course of the study.

Real-time ozone levels will be monitored to ensure they do not exceed the ACGIH threshold of 80 ppb. 80ppb. Upon completion of the study, an analyst-blinded regression will be conducted to determine differences in ozone between all rooms and arms.

### Tertiary Outcome 1: What is the characterization of antimicrobial resistance of ESKAPEE isolates, by trial site and sample matrix?

*Hypothesis*: ESKAPEE isolates in proximity of the exposed sinks exhibit elevated phenotypic resistance to antibiotics in comparison to ESKAPEE isolates from control sites.

*Approach*: Using the same samples and methods from primary objective 1 and secondary objective 1, selected presumptive ESKAPEE pathogens will be isolated on the same selective media. These isolates will be preserved and sent for whole genome sequencing to characterize any antimicrobial resistance in the genome. Additionally, Kirby-Bauer analysis will be conducted on the same isolates to identify the phenotypic resistance in parallel. All resistance profiles will be compared between isolates from the trial site and the sample matrix, and with available data from Bolivia and internationally_._

### Tertiary Outcome 2: Can we confirm phenotypic characterization of ESKAPEE pathogens via culture with whole genome sequencing of isolates?

*Hypothesis*: Culture methods for ESKAPEE pathogen detection and enumeration are reliable. *Approach*: Using the same samples and methods as in primary objective 1 and secondary objective 1, selected presumptive ESKAPEE pathogens will be isolated on the same selective media. Sequencing results will identify the taxa to the species level. Sequencing taxa assignment will be compared with our culture identification of the isolates to determine if there is a significant difference between phenotypic identification and genotypic confirmation.

### Study Methods

#### Sample size calculation

Sample size calculations for a cluster unmatched RCT with a binary primary outcome were conducted with baseline culture results. The primary outcomes for the baseline dataset and the intervention are the same: presence or absence of viable ESKAPEE pathogens. We used a binary outcome because our baseline data indicated that many sites had viable pathogens present, but counts were not consistently within a reliable range (30-300 CFUs). While CFU counts will still be collected, we will report the presence/absence of ESKAPEE pathogens as the primary outcome.

Baseline data from May 2025 were used to calculate the sample size. Only rooms where patients spend less than one hour per day were included. A cluster is defined by a sink area in which one 222nm lamp will be placed, around which a grouping of samples will be taken and pooled to count for one cluster sample, more on this method below. The binary outcome of positive/ negative for ESKAPEE pathogen was used, where the average positivity in Hospital A was 92.5%, Hospital B 60.3% and overall, 73.2% (Table 2). We ultimately calculate the *c*, number of clusters, and *n,* number of samples per cluster, to detect a statistically significant difference in the presence of ESKAPEE pathogens between the intervention and control group with 80% power at a 5% significance level. The calculations follow framework defined in Hayes & Bennett, 1999. First, we calculate the Intraclass Correlation Coefficient, _ρ_, with equation 1.

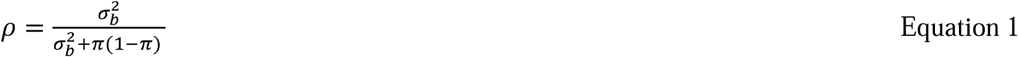

**Table 2.**
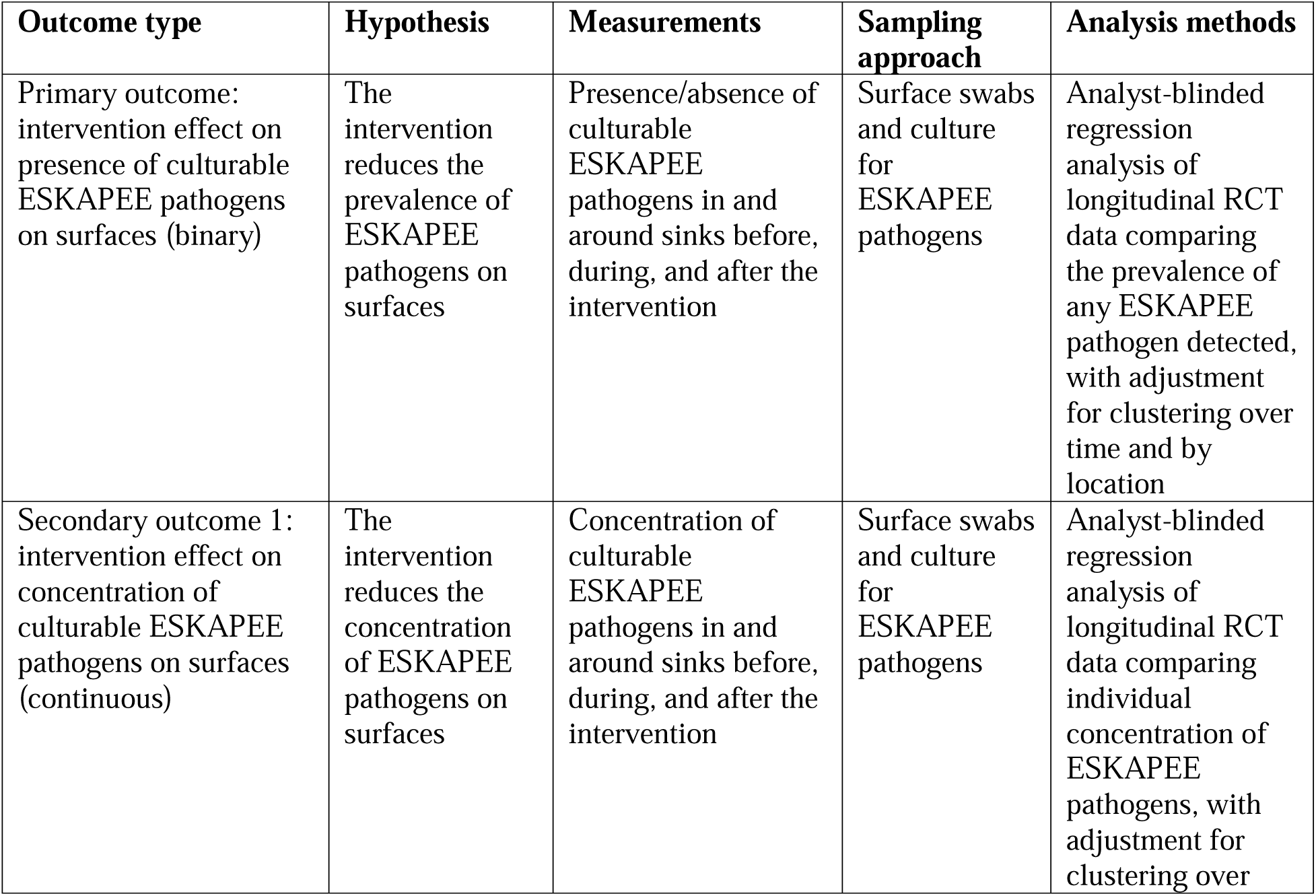
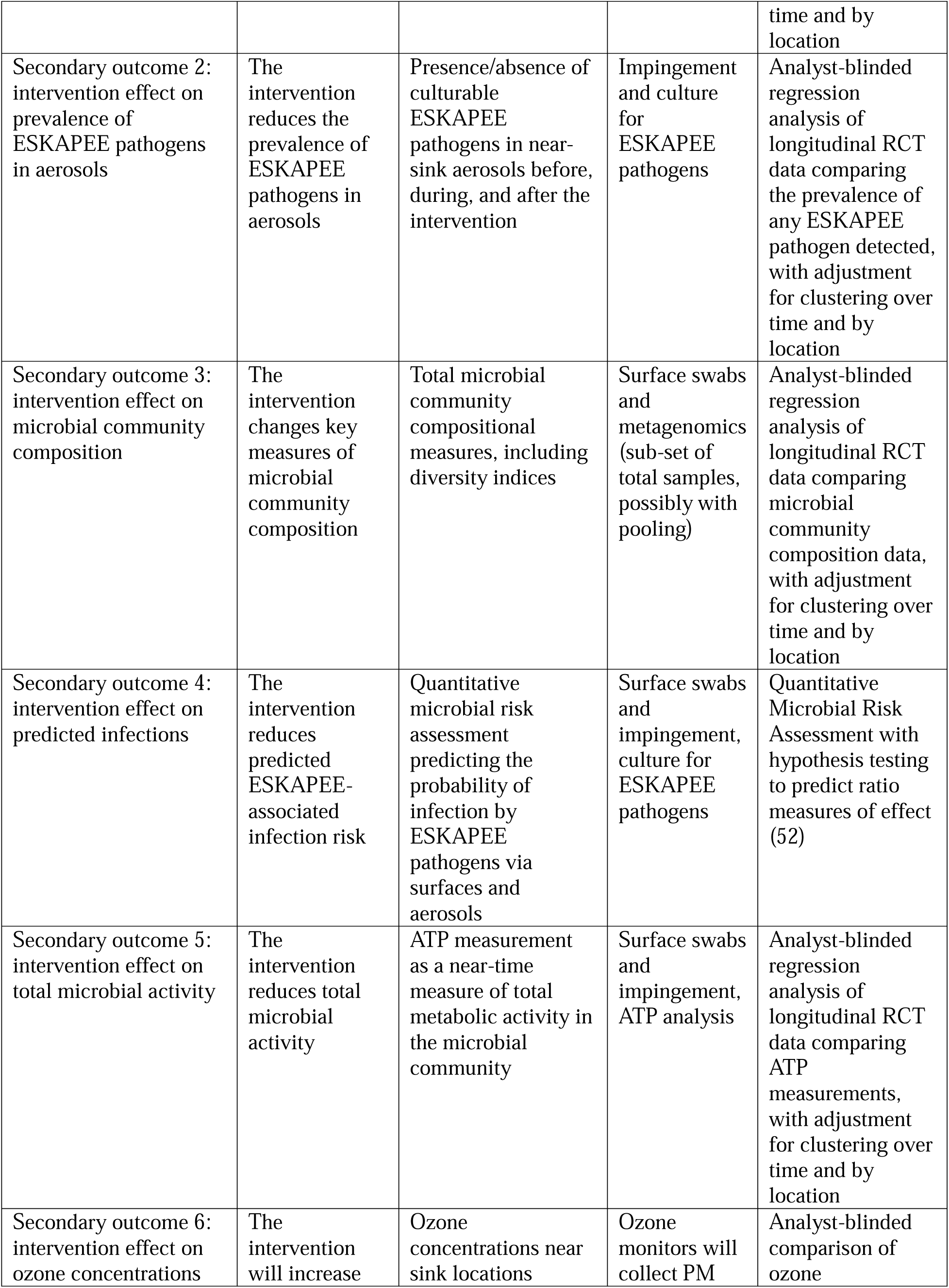
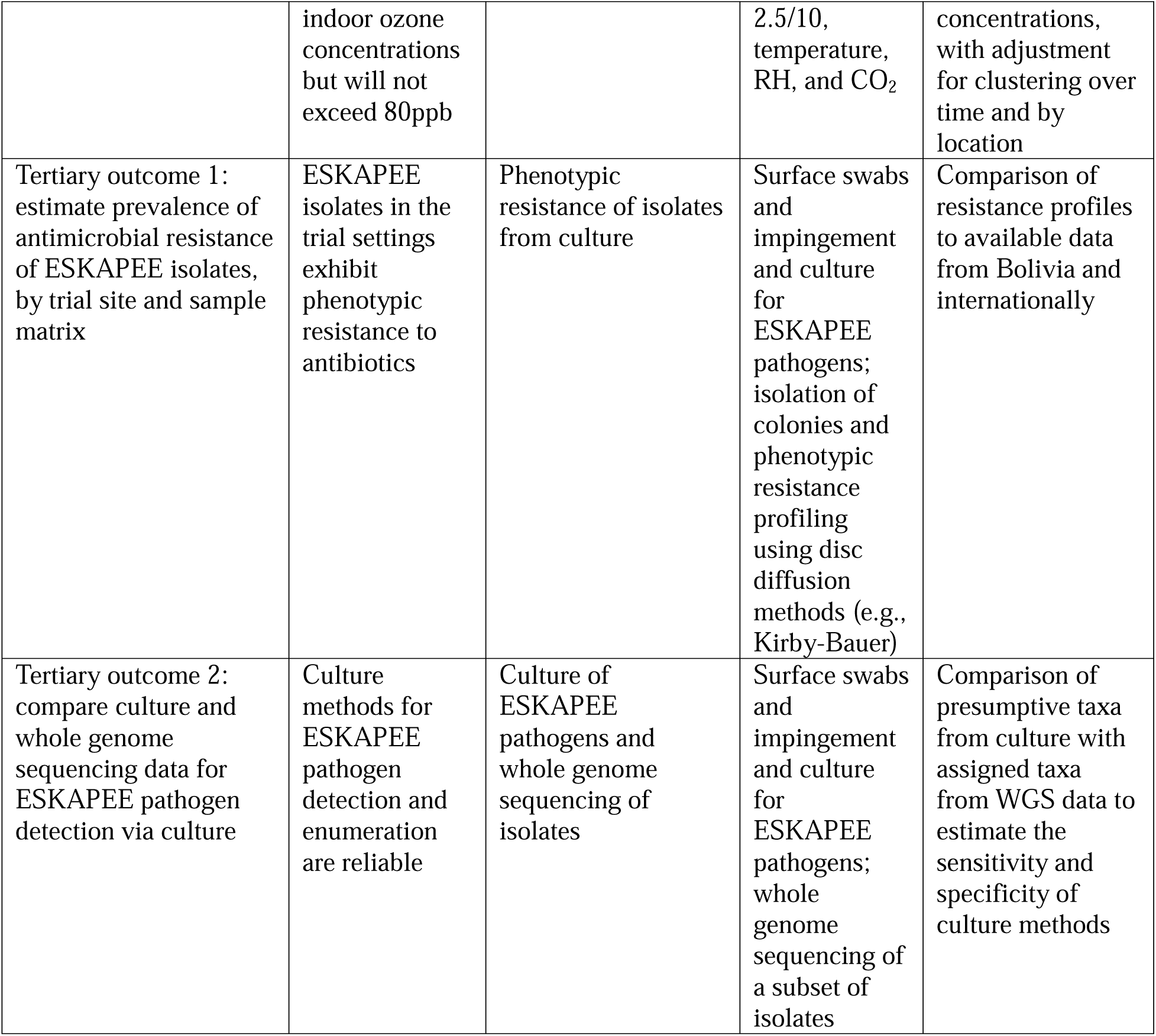
Study outcomes, hypotheses, sampling approaches, and analytical methods.

Where σ ^2^ is the variation between clusters, calculated as 0.088 based on empirical data, and π is the overall prevalence, calculated as 0.73. The overall _ρ_ was then calculated to be 0.33. We used the _ρ_ to estimate *k*, the coefficient of variation of true proportions between clusters within each arm of the study. Equation 2 reflects the conversion.

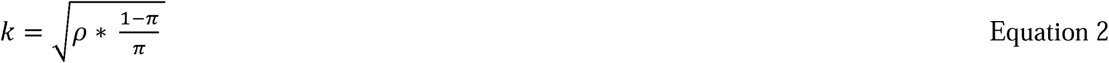

This calculation yields a *k* of 0.35. To determine the number of lamps per arm, *c*, and samples per lamp, *n*, for a study with power of 80% and 5% significance, we used equation 3.

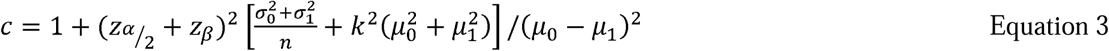

Where z_α/2_ and z_β_ are standard normal distribution values representing the upper tail probabilities at α/2 and β. Given the desired 80% power and 5% significance, these numbers were 1.96 and 0.84, respectively. The σ_1_ and σ_0_ are within-cluster standard deviations for the intervention and control arms, respectively. The mean prevalence of ESKAPEE pathogens in the control arm, μ_1_, stays constant at 0.73, while the intervention arm proportion, μ_0_, is variable based on our predicted detectable difference in ESKAPEE pathogen prevalence between the two arms. This calculation was applied to various possible scenarios, with a range in lamps per arm (*c*= 5,10,20,30,40,50) and plotted against the resulting *n* values which represent the number of lamps required per study arm in each hospital given the number of sampling points per lamp location. shows the number of lamps required per arm per hospital and the number of samples needed for a range of predicted detectable differences in ESKAPEE pathogens between the arms. Based on these calculations, at a ∼60% detectable difference, we plan to have 10 lamps per study arm per hospital and collect 10 samples at each sink location over the duration of the study. Each sampling event will yield one pooled sample from one cluster, therefore, 10 sampling events per sink will be necessary and we predict this to take 12 weeks.

**Figure 4.**
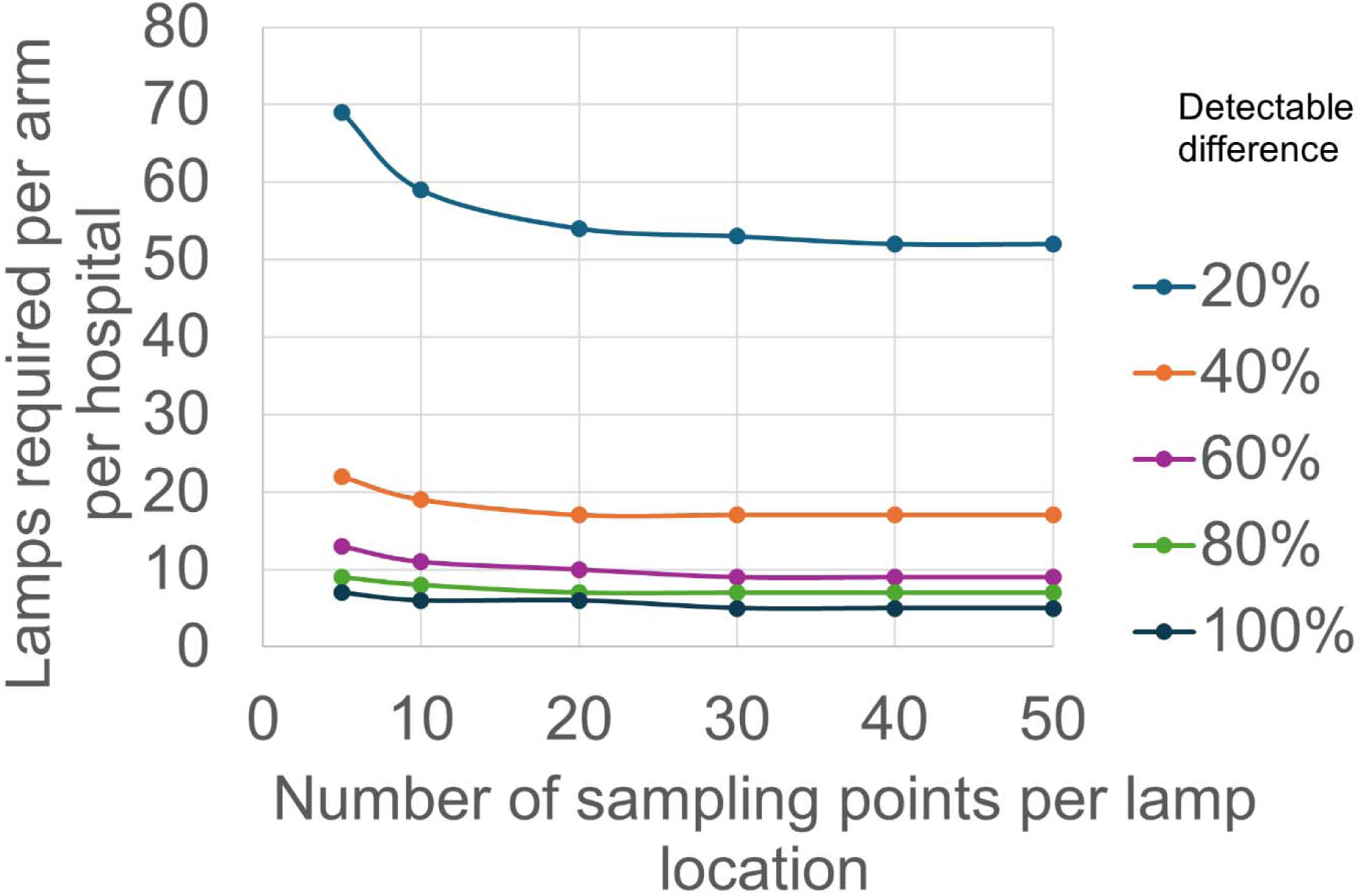
Number of sampling points per lamp location and number of lamps required per study arm per hospital for a range of detectable differences in ESKAPEE pathogens.

#### Cleaning protocol review

Prior to the start of the study, disinfection behaviors will try to be best understood by the study team. Cleaning protocols for all study sites will be collected by requesting any documentation hospital administration can provide. In addition to protocols, the study team will request cleaning schedules so that samples will be collected immediately before cleaning occurs. The goal is to sample as consistently as possible; directly before cleaning is scheduled. If changes or variations to protocols occur, the study team will adapt to sample as near to cleaning time as possible.

Additionally, cleaning logs to track which areas are cleaned how and when will be implemented a month before the intervention, and during. If there are changes in cleaning behavior, the logs will hopefully record them.

#### Selection of study sites

Two hospitals will participate in the study. Hospital A is a general hospital located adjacent to La Paz in a neighboring city, El Alto. This hospital is smaller with less resources. The other hospital, Hospital B, is located near the La Paz city center and is one of the most renowned hospitals in the county.

All sinks, also referred to as study sites, will be surveyed for inclusion and exclusion criteria, listed below. From this survey, a finalized list of eligible sinks areas will be compiled. The full survey for all details on the sinks, including measurements and usage, can be found in the supplemental information.

Inclusion Criteria:

Sink is used at least three times per day

There is sufficient counter and basin space to sample the necessary surface area

Exclusion Criteria:

Patients spend more than one hour per day in this room, unobstructed from the sink

Has a ceiling on which the Far-UVC lamp is not mountable

Non-hospital staff will be exposed to a lamp for more than 1 hour per day

#### Randomization and blinding

Assignment of study arm will be simple randomization and double-blinded to both the hospital staff and the sampling team. Each study site will be assigned an identifying number. A random generator will be used to assign 10 sites to the intervention arm and 10 to the control arm per hospital. These assignments will only be seen by a third-party professor at UMSA, who is familiar with the study design and hospitals, but is not involved in sample collection, processing, or analysis. This individual will be referred to as the blinder. They will write the assignments on three separate pieces of paper, which will be placed in three sealed envelopes and stored in three secure locations until unblinding. Figure 5 outlines the random assignment and blinding process. Blinding will continue until after the analysis is complete. Upon installation, the blinder will measure the irradiance on the surface of the counter. Over the course of the study, the blinder will measure and record the fluence weekly. If a functional issue with any UV lamp during the study cannot be resolved by the technical team, the lamp will be replaced with a new one.

**Figure 5.**
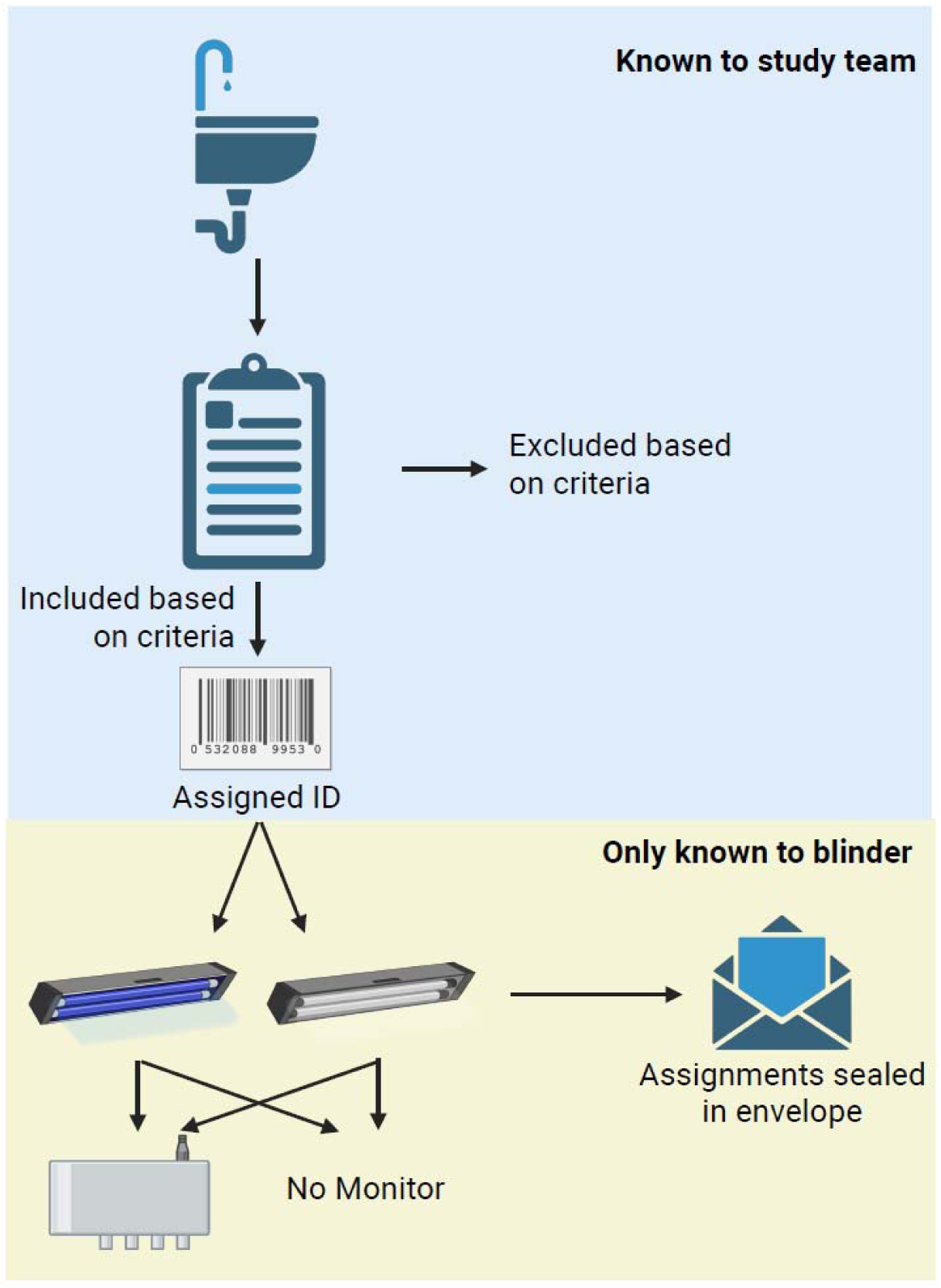
Random assignment process, with specific details for blinding.

#### Lighting design and installation

With the help of Karl Linden, A3 Lighting Consulting, and Ushio, each included study site will have a model built like Figure 1. Measurements taken from the inclusion/ exclusion room surveys (detailed in S1) will inform the placement of the lamps within the space. KrCL 222nm lamps (bulbs B1 Ushio Inc., fixtures UXV Inc.) will be installed directly above the sink. The highest UV dose will be directed at the sink basin. Official measurements will confirm the uniformity of sink and ceiling heights, which will inform us of adjustments of light intensity need to be made, to ensure uniformity among all sinks. A reasonable range of 15% difference in irradiation on the counter top is expected.

#### Running the lamps

Once the lamps are installed, all of them will be turned on within 30 minutes of each other. All lamps will run continuously, 24 hours per day, for the duration of the study (10 weeks). The lamps will run for 2 weeks before sampling begins. This will allow for microbial communities to normalize on sink surfaces from the lamps, and to combat the Hawthorne effect, when individuals alter their behavior because they are aware they are being observed. Sampling will begin after the two weeks are complete.

There are at times power supply disruptions. The power meters installed will be both to collect data on how much power the lamps use, and how frequently power interruptions occur. During the interviews with facility managers, we will also be asking questions about power disruptions, power supply, and priority wards to receive backup power.

### Sample/ data collection

#### Surface swabs

After the two-week waiting period, environmental sample collection will begin. Based on our sample size calculation, each study site must have 10 surface swab samples collected over the course of the study. A standard set of samples will be taken every sampling event that will include three surface swabs, outlined in figure 6. Surface swabs (Isohelix, Cell Projects Ltd. LLC) will first be wet with sterile Dey-Engley (D/E) Neutralizing Broth (Millipore), to reduce the effect of any recently used disinfectant products. Each of the three pre-determined sample locations will be swabbed using a 10 x 10 cm template to ensure identical areas. After swabbing is complete, the tip is broken off into 1 mL of D/E broth per swab and transferred on ice to the lab. Three field blank negative control swab samples will be collected over the sampling period to detect any field contamination. All 20 study sites will be sampled weekly, immediately before weekly or daily cleaning, for a total of 1,200 swab samples.

**Figure 6.**
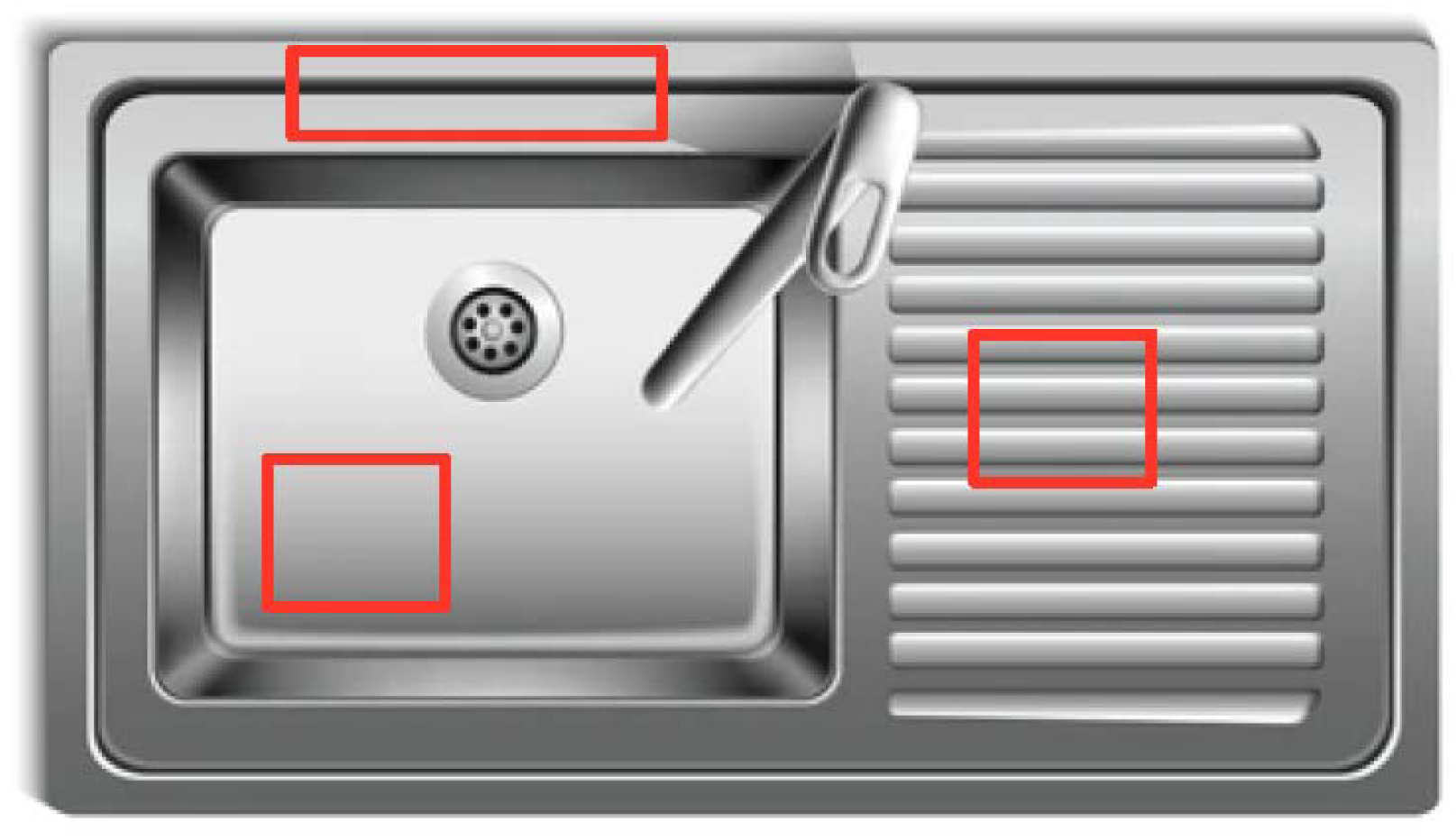
Swab sample plan. Highly contaminated areas according to baseline sampling and Kotay et al(58).

Upon arrival at the lab, tubes with swabs will be vortexed and the 1mL of buffer from all three samples pooled for an individual sink, for a combined 3mL (see Figure 6). One hundred uL of the combined solution will be plated on five different CHROMAgar selective media for ESKAPEE pathogens (Enterobacteria, ESBL, mSuperCARBA, Acinetobacter, Pseudomonas) and Tryptic Soy Agar as a general media. The remaining broth will be preserved with DNA/RNA shield and taken back to UNC for metagenomic sequencing. After 18-20 hours of incubation, colonies will be counted. Select colonies which are suspected to be ESKAPEE pathogens will be isolated and sent to UNC for whole genome sequencing. All samples and analysis methods are outlined in Table 3.

**Table 3.** Description and purpose of all samples to be collected throughout the study.

| Data | Sampling Method | Number of samples in intervention per hospital | Number of samples in control per hospital | Controls | Total number of samples | Analysis Method |
| --- | --- | --- | --- | --- | --- | --- |
| Surface-microbial | Surface Swabs | 3 per sink per week<br>10 sampling events<br>20 clusters<br>800 total | 3 per sink per week<br>10 sampling events<br>20 clusters<br>800 total |  | 1200 total swab samples<br>But when combined for plating, 400 total | Culture (Orientation, ESBL, Pseudomonas, Acinetobacter, S. aureus, TSA), TAC, sequencing |
| Surface-ATP | ATP fluorescence monitor | 10 per sink<br>20 clusters<br>200 total | 10 per sink<br>20 clusters<br>200 total |  | 400 | Luminultra bugcount |
| Air-microbial (Impinger/Air Cub) | Impingers, Air Cub | 3 per sink<br>20 clusters<br>60 total | 3 per sink<br>20 clusters<br>60 total |  | 120 | Culture (Orientation, ESBL, Pseudomonas, Acinetobacter, S. aureus, TSA), TAC, sequencing |
| Air-chemical | Ozone meters, Air quality monitors | 10 air quality<br>2 ozone | 10 air quality<br>2 ozone |  | 20 air quality<br>4 ozone | Comparative analysis |
| Cleaning log | Paper log implemented by hospital | 1 log for each hospital across all study sites |  |  |  | Comparative analysis of behavior between sites and before/ after intervention |
| Water usage | Ultrasonic flow meter | 5 flow meters | 5 flow meters |  | 10 flow meters | Count number of uses per day- try to equally distribute between arms and hospitals |
| Lamp/electricity usage | Power meter | 5 power meters | 5 power meters |  | 10 power meters | Count power disruptions or surges, usage cost- arm of study does not matter as much as equally distributed between hospitals |
| Lamp irradiance | Radiometer | Weekly by blinder | Weekly by blinder |  | 10 | Lamp functionality over time |

#### Air samples

Surface swabs will be taken before air sample collection begins. For the duration of the air sampling, the faucet will be on with cold water. The air sample will be taken with impingers. The machine will be placed within 0.5 meter of the sampling sink. The collection buffer will consist of 1% Peptone (0.2g), 0.01% Tween 80 (0.002g), and 0.05% Antifoam (0.01g), brought to 20 mL with sterile deionized water (56). The attached vacuum pump will be set to 28.3 Liters per minute and will run for 30 minutes(57). After 30 minutes, the collection buffer will be transferred into a sterile capped vile and transported back to the lab on ice. Over the course of the study, three air samples will be taken per study site, for a total of 120 air samples.

The impinger buffer will be directly plated (200uL) on the same six media mentioned above. After 18-20 hours of incubation, colonies will be counted. Select colonies which are suspected to be ESKAPEE pathogens will be isolated and sent to UNC for whole genome sequencing.

#### ATP swabs

ATP fluorescence data will be collected as a comparative method to the culture and molecular data. This is a rapid, real-time measure to semi-quantify the amount of ATP being produced from a surface swab. The method is typically used to determine the effectiveness of disinfection measures. Using the same template as the microbial swab collection, a swab will be taken and placed in the Lumiultra BugCount machine (Fuel Test Kit – 2nd Generation ATP). The hand-held machine will provide a real-time fluorescence measure, which will be recorded. One ATP sample will be taken at every site during each sampling day, for a total of 400 samples.

#### Chemical samples

For the duration of the study, several chemical monitors will be installed to monitor various compounds. At 20 study sites, a combined CO_2_, PM2.5/10, temperature, humidity, particle and formaldehyde monitor will be continuously collecting data (Temtop LKC-1000S+ 2^nd^). The monitors will be mounted 1.6 m from the floor, and the data will be stored internally on a memory card on the device. The monitors will be put in five intervention and five control sites in each of the hospitals.

Additionally, four ozone monitors will be mounted directly next to the CO_2_ monitors, two per study arm, to detect differences in overall ozone levels (2B tech, Personal Ozone Monitor, lower limit of detection of 3ppb).

Since the study is double blinded, the same randomization method will be used for allocation of the CO_2_ and ozone monitors; the sink IDs will be randomly selected to receive a monitor. The blinder will ensure equal distribution of the monitors across arms. All data will be stored on the respective monitors and collected at the end of the study. The research team will ensure that monitors are functional on a weekly basis, checking as they collect weekly microbial samples.

#### Functionality

Sink usage will be monitored in a subset of clusters, and an assumption will be made for all sinks from this measurement. Ultrasonic flow meters will be installed externally on 10 sink tail pipes, five in each hospital, to determine the number of times the sinks are used every day.

Power meters will be installed on 10 of the Far-UVC lamps, five in each hospital, to monitor power usage. This data will be used to confirm adherence to the 24-hour usage of the lamps, monitor for power outages or surges, and analysis of cost.

The blinder will visit each lamp on a weekly basis to measure and record the irradiance. This is to ensure the functionality of the lamps over time and to ensure the sample sites are receiving sufficient UV dosage. The blinder will share this data upon completion of the study.

### Primary Sample analysis

The selection of media is designed to gain a complete understanding of the presence and quantity of ESKAPEE pathogens in the samples. Though we can presume which taxa the colonies belong to, further molecular analysis is essential to confirm the species. Therefore, a select number of ESKAPEE pathogen will be isolated and sent for whole genome sequencing (WGS), while raw samples in D/E broth will be sent for metagenomic sequencing and qPCR (TaqMan Array) analysis. WGS will also give us more granular data on isolates, which will be examined for ARGs and compared to the culture and metagenomic data for similarities.

### Analysis Plan

All data, once collected, will be stored on personal computers of the study team, on a UNC OneDrive shared folder, and on Google Drive. Culture data will be uploaded and stored as it is collected. Chemical data will be stored in the internal memory of each monitor for the duration of the study and then uploaded in a single batch upon completion.

Our primary outcome is to compare the prevalence of ESKAPEE pathogens on sink surfaces in the control versus intervention arm using a regression analysis with Generalized Estimating Equations (GEE) to adjust for clustering. For our primary objective, a sample will be positive for ESKAPEE pathogen if any ESKPAEE pathogen grows on any media. We will then collapse the data to determine the percent of samples which were positive for ESKAPEE pathogen between the two arms. This will make the outcome continuous and a regression will be performed. For our secondary objective 1, we will be collecting continuous culture data, therefore we will have colony counts for each ESKAPEE pathogen on each media we plate, a similar regression will then be run.

We will compare the concentration of ESKAPEE pathogens in air between the control and intervention arms using a cluster-level unpaired two-sample t-test.

We will estimate the relative risk of infection from ESKAPEE pathogens between intervention and control arms using a quantitative microbial risk assessment approach. Briefly, we will employ a two-dimensional Monte Carlo simulation to estimate the risk of infection to ESKAPEE pathogens based on both sink contact and inhalation scenarios. We will compare the summative risk of infection between the intervention and control arms to estimate the relative risk and risk reduction attributed to the intervention at the sink level.

In addition to this analysis, metagenomics sequencing will assist in characterizing the entire microbiome of our samples. A regression analysis will be conducted to examine the differences between microbial communities amongst the study arms, hospitals, room function, etc.

Whole genome sequencing of ESKAPEE isolates will confirm identification and genotypic profiling, which will be compared to phenotypic profiling from Kirby Bauer testing. Antibiotics used for Kirby Bauer will include Vancomycin, Ampicillin, Linezolid, Gentamicin, Erythromycin, Ciprofloxacin, Oxacillin, Cefoxitin, Sulfamethoxazole/Trimethoprim, Clindamycin, Amikacin, Ceftriaxone, Ceftazidime, Meropenem, Amoxicillin/ Clavulanic Acid, Cefepime, Piperacillin/Tazobactam, Aztreonam, Gentamicin, and Cefotaxime.

We will compare the concentration of ozone between the control and intervention arms using a t-test. Room variations, occupancy, room use will be recorded from the baseline room survey, so these will be factored in (SI1).

All chemical and functional data will be monitored regularly throughout the study to ensure device functionality. During data analysis, trends and irregularities not detected over the course of the study will be compared with other variables to determine whether the results are affected.

### Communication Efforts

Upon completion of the study, the research team will first present to the hospital administration, then to any interested staff members in both hospitals. Presentations and a written summary will be provided to the hospital administration.

### Conclusion

This protocol outlines a Far-UVC effectiveness study to be implemented in an LMIC, the first of its kind. The intervention outlined is targeted at sinks, a known reservoir and amplifier of ESKAPEE pathogens(4, 5). Though some studies have sought to study the efficacy of Far-UVC, there have been a very limited number of studies which examine its effectiveness(59–63). This document aims to provide a wholistic summary of necessary baseline data, materials and methods, and analysis that will be conducted to conduct a large RCT, as proposed.

Based on our baseline data, 73.2% of all sink surfaces had culturable ESKAPEE pathogens. This is much higher than expected in High Income Countries(2). The baseline data, along with an expected detectable effect of 65%, makes for a samples size of 40 lamps, 10 lamps per arm of the study per hospital.

One limitation of this study design is that our primary outcome is binary rather than continuous. We determined the best measurable outcome is presence/ absence of ESKAPEE pathogens, and the continuous concentration being a secondary outcome of interest, due to non-predictable colony counts during baseline sampling.

## Data Availability

All data produced in the present work are available upon request to the authors.

## Acknowledgements

The authors gratefully acknowledge the hospital administrators, doctors, nurses, and cleaning staff for access to sampling sites and cooperation. This work was supported primarily by the Engineering Research Centers Program of the National Science Foundation under NSF Cooperative Agreement No. EEC-2133504.

