## Supplementary material for "The effects of Far-UVC irradiation on the presence and concentration of ESKAPEE pathogens on hospital surfaces: study protocol for a multi-site, double-blinded randomized controlled trial in La Paz, Bolivia": Suplemental Information

Appendix

Hospital Room Survey

To be administered at every potential sink

*Start of survey*

1. Hospital
   a. Hospital A
   b. Hospital B

*Location*

2. Room description/ name:
3. Floor number
   a. Fill in with number
4. Ward/ department type
   a. Fill in with text
5. Room usage
   a. Fill in with text
6. Is this a room where patients spend more than 8 hours ?
   a. Yes
   b. No
7. If yes- how many patients stationary for how long
8. If stationary (beds) – specify locations of beds and head location AND common activities of patients in bed for approx.. what times
9. Is this room ever used to provide pediatric care (care to patients under 18)
   a. Yes
   b. No
10. Is this room ever used for baby(?) deliveries?
    a. Yes
    b. No

*Measurements*

11. Distance between centroid of sink to centroid of likely light mounting location
    a. Fillin with number
12. How high is the ceiling
    a. Fill in with number
13. How high is the counter
    a. Fill in with number
14. Are there any objects or fixtures mounted above the sink (ex: cabinets) that will inhibit installation?
    a. Yes
       i. If yes, describe
    b. No
15. Any obstructions between the ceiling and the sink that will cast a shadow on the basin or counter top?
    a. Yes
       i. If yes, describe
    b. No
16. Location of sink in room
    a. Fill in with text
17. Length of sink (cm)
    a. Fill in with number
18. Width of sink (cm)
    a. Fill in with number
19. Height of sink (cm)
    a. Fill in with number
20. Counter length (cm) (if there is a counter around the sink)
    a. Fill in with number
21. Counter width (cm)
    a. Fill in with number
22. What is the ceiling made of?
    a. ceiling tile
    b. Plaster
    c. Dry wall
    d. Other (fill in blank)
23. How far from the sink (any side) is a light switch or outlet? (cm)
    a. Fill in with number
24. Photo of closest outlet/ switch
25. Is there a wire which is able to be split to power the lamp within 2m of the potential mounting location?
    a. Yes
    b. No
26. Based on your opinion, can a light be mounted above the sink?
    a. Yes
    b. No
       i. If no, why not?

*Usage*

27. Who uses this room?
    a. Doctors
       i. What do they do in the room?
       ii. How long do they spend in the room daily?
    b. Nurses
       i. What do they do in the room?
       ii. How long do they spend in the room daily?
    c. Patients
       i. What do they do in the room?
       ii. How long do they spend in the room daily?
    d. Cleaning staff
       i. What do they do in the room?
       ii. How long do they spend in the room daily?
    e. Visitors
       i. What do they do in the room?
       ii. How long do they spend in the room daily?
    f. Other (type in answer)
       i. What do they do in the room?
       ii. How long do they spend in the room daily?
28. Do any of the following demographics ever use this room?
    a. Children
    b. Immunocompromised individuals
29. Who uses this sink?
    a. Doctors
    b. Nurses
    c. Patients
    d. Cleaning staff
    e. Visitors
    f. Other (type in answer)
30. About how many times per hour is this sink used?
    a. Fill in with number
31. What is this sink used for? (choose any applicable)
    a. Hand washing
    b. Medical materials (ex: cleaning scalpels, tweezers, etc.)
       i. List medical materials
    c. Cleaning products (ex: bleach, hydrogen peroxide, etc)
       i. List cleaning products
    d. Dumping waste (ex: human waste)
       i. List waste
    e. Other (type in answer)
32. Other notes about usage
    a. Fill in text
33. Photos
    a. Attach photos

Cleaning protocols

34. Is there a cleaning protocol that applies to this room and sink areas?

Permanent people know the protocols, which are different for each area

Type of disinfection different and protocol

Some departments have logs but some don’t

When a new person comes in then they train the new people

a. Yes
b. No

If yes to 27

35. What level is this cleaning protocol defined at?
    a. Cleaning protocol specific to room
    b. Cleaning protocol specific to ward/unit
    c. Cleaning protocol is general across all patient care areas
36. Are there specific instructions for sink cleaning in the protocol?
    a. Yes
    b. No
37. Do cleaners receive formal training on the protocol
    a. Yes
    b. No
38. Is there a log to track when cleaning is performed?
    a. Yes
    b. No
39. Are there logs of verification (bacteria count etc.)?
